# Hospitalization Data Supports Correlation of Lower Covid-19 Severity vs. Universal BCG Immunization in the Early Phase of the Pandemic

**DOI:** 10.1101/2021.07.21.21260913

**Authors:** Serge Dolgikh

**Affiliations:** Dept. of Information Technology, National Aviation University, Kyiv

**Keywords:** Epidemiology, Covid-19, BCG, immunization

## Abstract

The possibility of a correlation between universal administration of the bacillus Calmette-Guerin (BCG) tuberculosis vaccine and lower severity of Covid-19 by national jurisdiction has been pointed out previously. In this work we examined hospitalization data attributed to Covid-19 cause reported by European national jurisdictions with the conclusion of a clear negative correlation between current or recent BCG vaccination program and reduced impact of the epidemics on the population measured in hospital admissions per capita in the early phase of the pandemic, before variants and vaccines. While there is no evidence that BCG vaccination provides strong individual level protection, the results of this work in combination with the results of other studies appear to support the hypothesis of a certain population-wide protection effect that is correlated with BCG immunization.

## 1 Introduction

Possibility of a correlation between universal bacillus Calmette-Guerin (BCG) tuberculosis vaccine program and lower initial impact of Covid-19 pandemic has been reported in a number of works previously [1–3]. Several studies reported statistically significant results [4,5] strongly supporting the correlation hypothesis, based on such factors as reported number of cases and mortality attributed to Covid-19.

One can consider the observable parameters such as: the number of hospitalizations with the main diagnosis of Covid-19; the number of cases with reported complications; the number of ICU admissions with the main diagnosis of Covid-19; and mortality with the main cause of Covid-19 as indicators of severity of the epidemics impact on the population, whereas the number of reported cases indicates the spread of the decease that is, its penetration in the population.

In this analysis we used openly available epidemiological data in the public domain to investigate the relation between the severity of the epidemiological impact expressed in hospitalizations per capita of population that were attributed to Covid-19 and the record of BCG immunization in the reporting jurisdiction, current or recent. The methods used are based on comparison, observation and include formal statistical analysis of the null hypothesis equivalent to absence of any correlation between universal BCG immunization program (UBIP) and the reported epidemiological impact in the jurisdiction.

## 2 Data Selection

### 2.1 National Hospital Care Statistics

A subset of cases of national hospital care statistics available in the public domain has been used to select a set of cases for the analysis. The selection was based on the following criteria:

- Availability of key statistics, such as cumulative and / or new hospital admissions attributed to Covid-19. New hospital admission statistics, such as weekly new admissions were converted to cumulative admissions in the period.
- Similarity with respect to background factors, including socio-economical, to reduce impact of secondary factors. Selected national cases are based in Europe and have similar socio-economic parameters.
- Variance with respect to UBIP factor: representative cases with respect to the factor of significance of the correlation hypothesis were selected.

The data in Table 1 was recorded in the initial phase of the pandemic, before variants and vaccines in the period Jan.1, 2020 to Aug.4, 2020 or from the first case reported in the jurisdiction. Data in the dataset was time-adjusted to approximately the same duration of the analyzed period after local arrival of the pandemics, of approximately 25 weeks. The epidemiological impact parameters, hospital admissions (*H*_*e*_) and ICU admissions (*U*_*e*_), where available, were measured per 100,000 capita of the population. UBIP record (last column) is measured by group factor, A: current UBIP program; B: previous UBIP, year of cessation; C: no previous universal BCG vaccination program or equivalent. Refer to [5,6] for further details.

**Table 1.** National hospital care statistics dataset, Europe and North America

| *Case* | *Population, M* | *Local arrival* | *Hospital adm.* | *ICU adm.* | *UBIP record* |
| --- | --- | --- | --- | --- | --- |
| Greece | 10.5 | 28.02 | 17.60 | 3.05 | A |
| Finland | 5.5 | 29.01 | 25.0<sup>(1)</sup> | - | B, 2006 |
| Norway | 5.4 | 26.02 | 20.42 | 4.36 | B (1995) |
| Germany | 82.8 | 27.01 | 29.71 | - | B (1998) |
| Portugal | 10.3 | 02.03 | 57.03 | - | A |
| Ireland | 4.9 | 29.02 | 41.43 | 5.3 | A |
| USA | 327.2 | 21.01 | 97.41<sup>(2)</sup> | 4.48<sup>(2)</sup> | C |
| Netherlands | 17.2 | 27.02 | 65.51 | 16.61 | C |
| Italy | 60.5 | 31.01 | 151.24 | - | C |
| Spain <sup>(3)</sup> | 46.7 | 31.01 | 242.70 | 20.83 | B2 (1965-81) |
| UK <sup>(4)</sup> | 66.4 | 29.01 | 120.61 | - | B3 (2007) |
| Belgium | 11.4 | 04.02 | 172.03 | - | C |

<sup>(1)</sup> Estimated
<sup>(2)</sup> Likely underreported, as not all states reported consistent statistics [7]
<sup>(3)</sup> Considered group C equivalent due to short duration of UBIP
<sup>(4)</sup> Considered group C equivalent due to age administration practice incompatible with the early-age induced protection hypothesis, [2]
**Sources:**
European Center for Disease Control hospital care statistics [8]
US Covid Data Tracker [7]
Our World in Data: Covid-19 Hospitalizations [9]; other sources [10-14]

## 3 Results

### 3.1 Observations

A histogram of the cases in the dataset by the epidemiological factor *H*_*e*_ of hospital admissions is shown in Figure 1.

**Fig.1.**
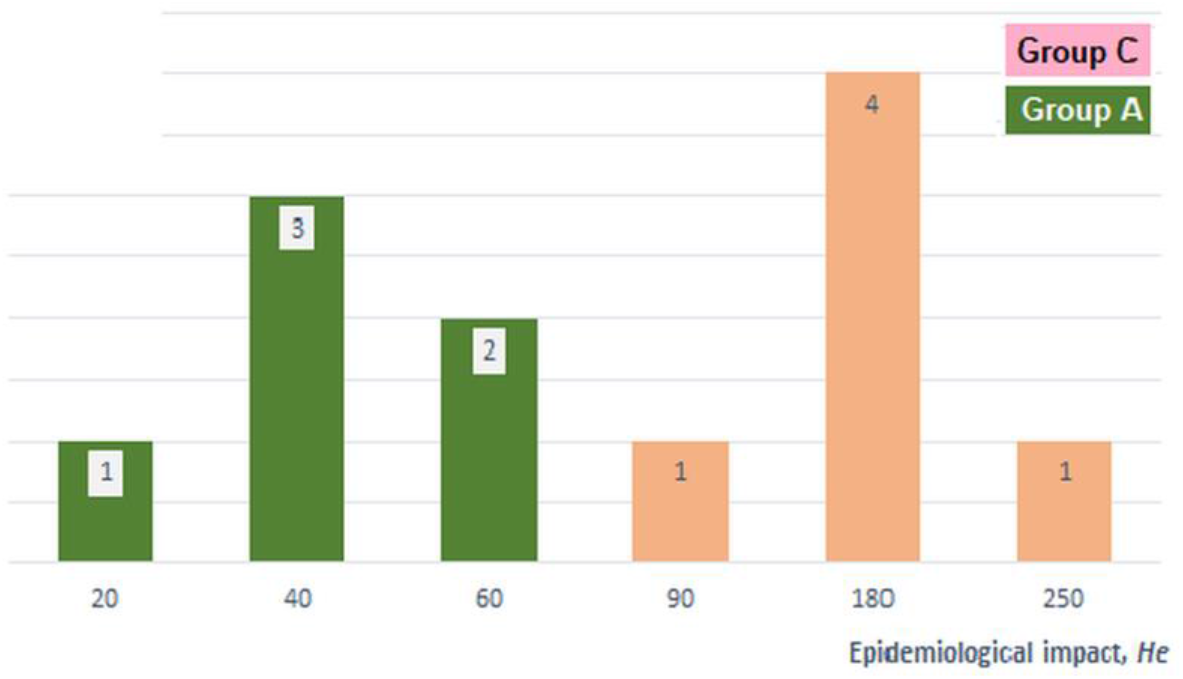
Epidemiological impact *H*_*e*_ by UBIP group.

Grouping of cases can be observed with clear correlation with the UBIP group. Specifically, the maximum value of the impact factor in the positive UBIP group of cases (A, B) was lower than the minimum value in the negative group (C and equivalent), and the mean of the positive group was significantly lower, in the observed epidemiological impact, than that of the negative group:

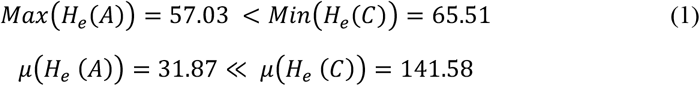

A numerical value in the interval [0, 1] can be assigned to the UBIP factor *U*_*f*_ (Table 1), with the value of 1 signifying group A (current UBIP), that of 0, group C and for cases in group B, a value between 0 and 1 depending on the time of cessation of UBIP. Calculation of the correlation coefficient between the factors *H*_*e*_ and *U*_*f*_ produced strong negative value of −*0*.*809*, indicating that stronger immunization record was strongly correlated, in the analyzed period and dataset, with the lower recorded epidemiological impact.

A direct observation of the histogram of the dataset by reported epidemiological impact in Figure 1 confirms that it has an appearance of a strongly expressed bimodal distribution with significantly different means (1), in clear contradiction to the null hypothesis.

Similar pattern can be seen with a different observable parameter associated to severity of Covid-19 scenario: ICU admissions *I*_*e*_ though it was not available for all jurisdictions in the dataset resulting in a smaller set of statistics.

### 3.2 Statistical Significance Analysis

The statistical significance analysis is based on an observation that under the null hypothesis, the groups identified by UBIP record, namely, groups A and C represent samples of a given size from a presumably normal distribution with parameters *μ, σ* randomly drawn, as *U*_*f*_ under the null hypothesis, would have no correlation with the epidemiological outcome, *H*_*e*_. The distribution of sample means then is governed by the central limit theorem, that allows to place constraints on the p-value of the null hypothesis under the observed parameters of the group samples.

By applying the analysis described in [5] to the hospitalization statistics dataset in Table 1, one obtains:

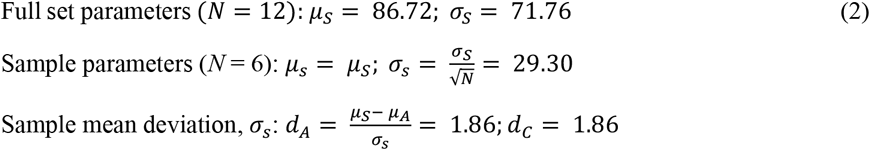

The p-value of the null hypothesis is then obtained as the product of independent, under the null hypothesis, probabilities of deviations of sample means *p(d*_*A*_) and *p(d*_*C*_) from the set mean:

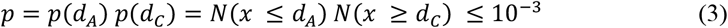

From (3) it follows that with the dataset and distribution of sample parameters in the analysis, the null hypothesis is excluded at the confidence level of at least 0.001, in agreement with earlier statistical results based on different epidemiological impact parameters [4,5].

## 4 Possible Mechanisms

Observing the course of epidemiological scenarios in different jurisdictions does not provide sufficient grounds to expect that BCG immunization could offer strong individual level protection against Covid-19, and possibly other infections and conditions [15], similar to that of specific vaccines. Rather, the observed scenarios can be associated with mitigated or attenuated infection resulting in lower and perhaps, less severe initial impact on the population.

While studies are ongoing to narrow down possible specific mechanisms and channels of such broad and general protection, it can be hypothesized that formation of some general immune mechanism can be triggered by an exposure to attenuated tuberculosis bacilli in the early age, resulting in a more rapid and effective immune response to an exposure to a broad range of infections of respiratory and other types in significant part of the immunized population. Possible effects of such type of pretrained immunity can be, for example, lower production of viral load and avoidance of strong auto-immune reaction, with a possibility of multiple further effects, including reduced time of infectivity, reduced viral load in transmission and lighter course of the disease, all contributing to the overall pattern of “dampening” or slowing the rate of the development of the infection in the population.

## 5 Conclusions

Several observable parameters such as: the number and rate of hospitalizations; that of the reported complications; ICU admissions; and mortality attributed to Covid-19 as primary cause, can be associated with the severity of the epidemiological scenario in a reported jurisdiction. A number of previously reported results based on statistics of the reported cases and mortality pointed to a possibility of correlation between severity of Covid-19 scenario and universal BCG immunization program in the jurisdiction. In this work, the same conclusion is supported by an analysis of hospitalization statistics in set of European jurisdictions with similar socio-economic parameters. The consistency of the finding with a different statistic in our view, strengthens the case for the correlation hypothesis.

Another conclusion of this study is that availability of diverse, compatible and comprehensive statistics in the open access can have clear and strong benefit for the research into the nature of the epidemics and approaches that would be able to mitigate its impacts on the society. From that perspective, availability of statistics of reported complications, ICU admissions and other in a consistent format across broader set of jurisdictions, national as well as subnational, regional, etc., would allow to perform more comprehensive statistical analysis of the hypothesized correlation and improve its confidence.

Epidemiological scenarios observed worldwide indicate that even if the causation of less severe epidemiological scenario can be confirmed, BCG immunization may not offer strong society-wide resistance to general respiratory viral infection. Rather, it may mitigate its impacts temporarily, perhaps via one of the discussed channels providing an important window for development of more effective measures and policies.

In conclusion, it needs to be emphasized that the results and conclusions of this analysis apply to the early phase of the pandemic, before the infection had a chance to mutate significantly and the influence of other factors, for example, policy decisions, vaccinations, could impact the course of the epidemiological scenario to a significant extent.

## Data Availability

Data referred to in the manuscript is openly available without registration

https://www.ecdc.europa.eu/en/publicationsdata/download-data-hospital-and-icu-admission-rates-and-current-occupancy-covid-19

https://covidtracking.com/data/national/hospitalization/

https://ourworldindata.org/covid-hospitalizations

http://www.bcgatlas.org/

https://www.worldometers.info/world-population/

https://thl.fi/en/web/infectious-diseases-and-vaccinations/what-s-new/coronavirus-covid19-latest-updates/situation-update-on-coronavirus

